# Global research on syndemics: A meta-knowledge analysis (2001-2020)

**DOI:** 10.1101/2021.05.19.21257413

**Authors:** Md Mahbub Hossain, Nobonita Saha, Tahmina Tasnim Rodela, Samia Tasnim, Tasmiah Nuzhath, Tamal Joyti Roy, James N. Burdine, Helal Uddin Ahmed, E. Lisako J. McKyer, Banga Kamal Basu, Ping Ma

**Affiliations:** Texas A&M University; University of Dhaka; Mawlana Bhashani Science and Technology University; Khulna University of Engineering and Technology; National Institute of Mental Health; Gazi Medical College

**Keywords:** Syndemics, Multimorbidity, Health inequity, Health disparity, Health Promotion, Health Policy, Global Health, Meta-knowledge Analysis

## Abstract

Syndemics or synergies of cooccurring epidemics are widely studies across health and social sciences in recent years. We conducted a meta-knowledge analysis of articles published between 2001 to 2020 in this growing field of academic scholarship. We found a total of 830 articles authored by 3025 authors, mostly from high-income countries. Publications on syndemics are gradually increasing since 2003, with rapid development in 2013. Each article was cited more than 15 times on average, whereas most (n = 604) articles were original studies. Syndemics research focused on several areas, including HIV/AIDS, substance abuse, mental health, gender minority stressors, racism, violence, chronic physical and mental disorders, food insecurity, social determinants of health, and COVID-19. Moreover, biopsychosocial interactions between multiple health problems were studied across medical, anthropological, public health, and other disciplines of science. The limited yet rapidly evolving literature on syndemics informs transdisciplinary interests to understand complex coexisting health challenges in the context of systematic exclusion and structural violence in vulnerable populations. The findings also suggest applications of syndemic theory to evaluate clinical and public health problems, examine the socioecological dynamics of factors influencing health and wellbeing, and use the insights to alleviate health inequities in the intersections of synergistic epidemics and persistent contextual challenges for population health.

## 1. Introduction

Addressing health inequities in marginalized populations requires a complex understanding of the epidemiological burden of multiple health problems and associated factors that determine the health statuses and outcomes in that context [1–3]. Theoretical frameworks may help in examining different dimensions of population health, adopting intellectual inspirations from scholarly concepts that emerged in the past across scientific disciplines [4, 5]. Amongst many contemporary theories, “syndemic(s) theory” offers critical perspectives on the relationships between diseases and biopsychosocial factors that not only explain the high burden of diseases in populations but also sustain a series of adverse health and social outcomes [6, 7]. Conceptually, syndemics have three fundamental components that characterize them [6, 8, 9]. Firstly, two or more health problems cluster together that can be assessed epidemiologically or described as co-morbidity or multimorbidity. Secondly, syndemic diseases or conditions interact among themselves using biological, psychological, or social pathways. Lastly, syndemics are associated with social, structural, and contextual forces that precipitate disease clustering and progression in the first place, which may include but not limit to poverty, segregated housing, systematic exclusion from opportunities, enslavement, colonialism, neo-colonialism, and neo-liberal economic measures that result in disproportionate distribution of wealth, lack of access to resources and services that may improve health and wellbeing, and other socioeconomic inequalities [6, 8–10].

Since its conceptualization by Merrill Singer in the 1990s [10–12], syndemic(s) theory has become one of the eminent ideas that has influenced scholarly discourses in different disciplines, including social epidemiology and medical anthropology, investigating the dynamics of persistent health inequities in disadvantaged population groups such as homeless, racial and ethnic minorities, and people affected by chronic diseases or social problems [6, 9]. One of the earliest examples used in theorizing syndemics was substance abuse, violence, and HIV/AIDS (SAVA) [8, 10]. The case of the SAVA syndemic illustrated the clustering of these three health problems where a high burden of substance abuse and violence were prevalent in people living with HIV/AIDS, predominantly in the systematically oppressed communities in the United States [10]. Most of them were urban poor, people of color, and deprived of opportunities to live healthy lives in the first place. Another syndemic of HIV and Hepatitis C virus (HCV) is recognized in people who use drugs (PWUD) [9, 13]. Nearly 2.3 million out of 36.7 million people living with HIV in 2015 were infected with HCV [14]. The co-infection of HIV and HCV significantly increases the risks of advanced liver disease and associated adverse health outcomes compared to those with HCV infection alone [9, 15, 16]. Moreover, HCV facilitates the pathogenesis and disease progression of HIV, thus interacting with each other in multiple pathways that may result in adverse health outcomes among the affected individuals [16]. These infections are highly prevalent in PWUD, who are more likely to share syringes and engage in other high-risk health behaviors. In addition to these pathological and psychological challenges, these people share similar socioeconomic marginalization driven by structural forces that transform their life choices, health behaviors, and biopsychosocial outcomes [9, 16]. Rather than presenting the disease burden and their correlates only, syndemic(s) theory highlights the complicated relationships between these co-existing health problems that can be biological, psychological, or social in nature. More importantly, studying these problems in the context of their shared determinants and interactions between multiple constructs provides a broader understanding to address the problems, which is less probable if these problems are examined individually without exploring their synergistic characteristics [9, 13].

A growing body of literature indicates that the academic and professional interests in syndemics have increased over the past three decades [6, 7, 9]. A review of syndemics associated with HIV/AIDS identified 60 articles published in 2019 alone [6], which reported co-conditions such as substance abuse (n = 40; 67%), high-risk sexual behavior (n = 36; 60%), depression (n = 36; 60%), interpersonal violence (n = 35; 58%), stigma (n = 19; 32%), sexually transmitted infections (STIs) (n = 16; 27%), trauma (n = 14; 23%), and noncommunicable diseases (NCDs) (n = 6; 10%). Such inclusive nature of those syndemics literature informs the scope of interdisciplinary research using tools from diverse sources to answer common questions of interest in different contexts and populations. The increasing recognition of research on syndemics can potentially specify, describe, and explain bio-social interactive pathways advancing both science and practice. Another review identified 143 journal articles, 23 book chapters, and 29 other types of publications [9]. In this review, the authors reported five thematic categories of studies that included syndemics (12%) with cooccurring diseases with bib-bio and bio-social interactions, potential syndemics (18%) where those interactions are referred but not fully articulated, socially determined heightened health burden of diseases (15%) that described social conditions associated with poor health without identifying disease clusters or interactions, harmful disease clusters (17%) that identified disease clusters and social factors without mentioning their interactions, and adverse additive co-morbidities (38%) describing diseases and social determinants through an additive approach to adverse health outcomes without examining the evidence on interactions. Furthermore, a systematic literature review assessed a potential syndemic comprising of HIV, HCV, intimate partner violence, and posttraumatic stress disorder (PTSD) [17]. In this review, the authors reported childhood physical and sexual abuse and intimate partner violence as social sources of trauma, which was associated with elevated unsafe health behavior leading to a higher burden of HIV infections and subsequent health outcomes. Existing literature provides an overview of diverse health problems and methodological measures adopted by different authors to study syndemics and associated conditions [6, 9], which highlight both the complexity of the current evidence base and the necessity of improving our understandings of syndemics applied in different frontiers of knowledge.

Primary studies and analytical reviews offer syntheses of evidence focusing on specific research questions relevant to a field or a problem of interest, which are useful for evaluating evidence in that scenario. However, such focused approaches may not provide a comprehensive overview of the entire scientific landscape or describe how research on a domain or topic evolved over time. In this regard, meta-knowledge analyses offer quantitative assessments of research identifying the overall status and characteristics of research. Through scientometric and bibliometric measures, meta-knowledge studies identify top contributing scholars, institutions, journals, and countries that may promote further research collaborations. In addition, meta-knowledge studies aim to identify research hotspots where most studies have emphasized previously, thus inform research trends and explore research areas that are not examined extensively. Recognizing areas that require further studies is one of the many goals of knowledge development in a field or topic of interest. To the best of our knowledge, there is no meta-level study on syndemics, which informs an overview of the current status and the evolution of global knowledge on syndemics-related research. Such studies, if conducted systematically, can inform the scholars and practitioners to understand the historical development of knowledge in this field, explain the status of intellectual advancements, and guide future research and scientific advancements in this area of growing interest in health sciences research. In the current study, we primarily aimed to analyze the characteristics and trends of the global research literature on syndemics. Secondarily, we evaluated the most prolific authors, institutions, journals, affiliating countries, and funding institutions contributing to syndemics-related research. Lastly, we mapped the major knowledge domains on syndemics highlighting the intellectual development in this field.

## 2. Materials and methods

### 2.1 Data source

In this study, we adopted meta-knowledge methods that have been used in previous research [18, 19]. To retrieve scientometric data on syndemics, we accessed the Web of Science (WoS) core collection that included multiple citation sources such as Science Citation Index-Expanded (SCI-Expanded), the Social Sciences Citation Index (SSCI), the Arts & Humanities Citation Index (A&HCI), and the Emerging Sources Citation Index (ESCI). The selection of WoS as the data source was informed by several advantages that it offers. Firstly, WoS provides extensive coverage of more than 20,000 journals, making it one of the most widely used databases for bibliometric studies [20]. Moreover, WoS includes publications not only from biomedical sources but also social sciences and other scholarly disciplines, thus making the bibliographic collective more inclusive in nature [21, 22]. As syndemics are associated with a wide range of biopsychosocial issues relevant to different scientific disciplines [9, 23], WoS is likely to provide a transdisciplinary overview of the research landscape in this topic.

### 2.2 Data extraction

We used “syndemic*” keyword in the topic field for searching the titles, abstracts, and keywords in the WoS database collection. The search process was structured using several eligibility criteria. First, we included citations published from January 1, 2000, to December 31, 2020. Second, we reviewed the titles and abstracts of the retrieved citations and excluded studies on topics such as “parasyndemicolpate” OR “syndemicolpate” that could have provided false positive entries. Third, we limited our search in citations published in English language, thus we excluded citations published in languages other than English. Lastly, we included all publication types such as original articles, reviews, commentaries, editorials, letters, and book chapters considering a low number of citations in the emerging field of knowledge. For all eligible citations, the complete bibliometric data on publication records, authorship, institutional affiliations, funding information, keywords, and citations data were extracted for subsequent analytic steps.

### 2.3 Data analysis

The corpus of the eligible literature was used for descriptive analysis, social network analysis, and conceptual structure analysis using measures that have been used in previous knowledge mapping studies [18, 20, 24, 25]. The descriptive analyses on the key bibliometric characteristics such as total citations, publication trends, top ten cited articles, prolific authors, h-index and g-index of the authors, contributing journals, research institutions, affiliating countries of the authors were analyzed using *Microsoft Excel* [26] and *R* software (version 4.0.2) [27, 28].

Knowledge mapping offers visual representations of the social connectedness among affiliating authors, institutions, and countries that reflect the research collaborations in a topic. We used the *VOSviewer* software that applies a natural language processing algorithm (NLP), where the units of analysis are represented within the maps as circular nodes [29]. The size of the node accounts for volume (e.g., number of publications in a dataset) and the position represents the similarity with other nodes in the same set. In this mapping process, closer nodes are more alike than nodes far apart from each other. The lines connecting multiple nodes represent the relationships among those nodes, whereas the thickness of those lines indicates the strength of that relationship. Finally, the color of the node indicates the cluster to which each node has been allocated to. In this way, all nodes in the map are clustered together based on their relatedness (Van Eck et al., 2010). To make the map, *VOSviewer* uses the SMACOF algorithm [30], which minimizes the function:

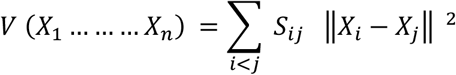

under the constraints:

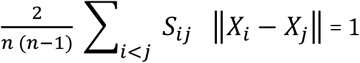

where:
n–the number of nodes in a given network,

X_*i*_–the locations of node *i* within a two-dimensional space,

∥X_*i*_-X_*j*_∥–the Euclidean distance between nodes *i* and *j*.

*VOSviewer* builds clusters of nodes by maximizing the following function:

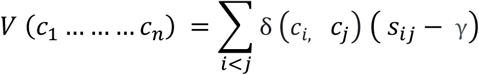

where:
ci–the cluster to which node *i* is assigned,

δ (c1, cj)–a function that equals one if ci = cj; and zero otherwise,

γ–a resolution parameter that determines the level of detail of the clustering (the higher γ is, the higher the number of clusters).

This method uses a distance-based approach to construct the bibliometric maps in three steps [29, 31]. At the first step, it normalizes the differences between multiple nodes. At the next step, it builds a two-dimensional map where the distance between multiple nodes reflects the similarities between those nodes. Further, it combines closely related nodes into clusters sharing similar bibliometric properties allowing visual representation of the research field. Moreover, we conducted a co-occurrence analysis of keywords to evaluate the conceptual relationships between multiple topics. The frequency of co-occurrence of two or more keywords indicated the strength of their association, whereas multiple keywords appearing within a cluster highlighted the topical foci or knowledge base within the research landscape. This approach was used to map multiple clusters with an overview of research domains and a graphical evolution of those domains across years. Lastly, we developed a three-field plot connecting the current literature with cited references using *KeyWords Plus* in the WoS database. This was used to generate the most frequently appearing words or phrases in cited sources, depicting the intellectual linkages between the existing studies on syndemics and cited research articles. This meta-knowledge study is registered in the Open Science Framework, and the data on eligible studies can be found in the following repository (https://osf.io/b4mvr/).

## 3. Results

### 3.1 Overview of the publications

We found a total of 830 articles eligible for this bibliometric study, including 604 original articles, 75 reviews, and 151 other types of publications from 314 scholarly sources (Table 1). These documents were authored by 3025 individual authors, with more than three authors per document. Most (n = 750) articles had multiple authors, whereas 80 articles were authored by a single author. The collaboration index was 4.03 suggesting more than four co-authors per article index calculated only using the multi-authored article set.

**Table 1:** Summary of the bibliographic findings.

| **Description** | **Frequency** |
| --- | --- |
| No. of total documents | 830 |
| No. of scholarly sources | 314 |
| No. of original articles | 604 |
| No. of review articles | 75 |
| No. of other publications (letters, editorial etc.) | 151 |
| No. of author's keywords | 1492 |
| No. of authors | 3025 |
| No. of single authored documents | 80 |
| No. of cited references | 30189 |
| Mean citations per document | 15.09 |
| Mean annual citations per document | 2.35 |
| Mean authors per documents | 3.64 |
| Mean documents per author | 0.274 |
| Collaboration index | 4.03 |

Table 2 provides an overview of the top 10 cited articles on syndemics published from 2003 to 2019, with the average citation per article per year ranging from 12.81 to 124.67. Seven of these articles focused on HIV-related syndemics, whereas the remaining articles discussed HIV in syndemic-related discourses emphasizing other health issues such as substance abuse, tuberculosis, violence, nutritional disorders, chronic diseases, and social determinants of health.

**Table 2:** Top ten cited articles on syndemics research.

| **Title** | **Authors** | **Journals** | **Publication Year** | **Total Citations** | **Average Citations per Year** |
| --- | --- | --- | --- | --- | --- |
| Syndemics and public health: Reconceptualizing disease in bio-social context | Singer, M; Clair, S | Medical Anthropology Quarterly | 2003 | 480 | 25.26 |
| The Global Syndemic of Obesity, Undernutrition, and Climate Change: The Lancet Commission report | Swinburn, B A. et al. | Lancet | 2019 | 374 | 124.67 |
| Psychosocial health problems increase risk for HIV among urban young men who have sex with men: preliminary evidence of a syndemic in need of attention | Mustanski, B. et al. | Annals of Behavioral Medicine | 2007 | 328 | 21.87 |
| HIV and Tuberculosis: a Deadly Human Syndemic | Kwan, C.K.; Ernst, J.D. | Clinical Microbiology Reviews | 2011 | 310 | 28.18 |
| Antiretroviral Therapy for Prevention of Tuberculosis in Adults with HIV: A Systematic Review and Meta-Analysis | Suthar, A.B. | Plos Medicine | 2012 | 233 | 23.3 |
| Syndemics and the biosocial conception of health | Singer, M. et al. | Lancet | 2017 | 229 | 45.8 |
| Sexual Compulsivity, Co-Occurring Psychosocial Health Problems, and HIV Risk Among Gay and Bisexual Men: Further Evidence of a Syndemic | Parsons, J.T. et al | American Journal of Public Health | 2012 | 206 | 20.6 |
| Syndemics, sex and the city: Understanding sexually transmitted diseases in social and cultural context | Singer, M. et al. | Social Science & Medicine | 2006 | 205 | 12.81 |
| Syndemic Theory and HIV-Related Risk Among Young Transgender Women: The Role of Multiple, Co-Occurring Health Problems and Social Marginalization | Brennan, J. et al. | American Journal of Public Health | 2012 | 199 | 19.9 |
| Substance Abuse, Violence, and HIV in Women: A Literature Review of the Syndemic | Meyer, J. et al. | Journal of Women's Health | 2011 | 178 | 16.18 |

### 3.2 Publication trends

Figure 1 highlights an increasing trend of scholarly publications on syndemics since 2003. Only 33 articles were published until 2010, whereas the frequency of publication increased significantly since 2013. The annual growth rate of the scholarly publications on syndemics was calculated as 10.47%.

**Figure 1:**
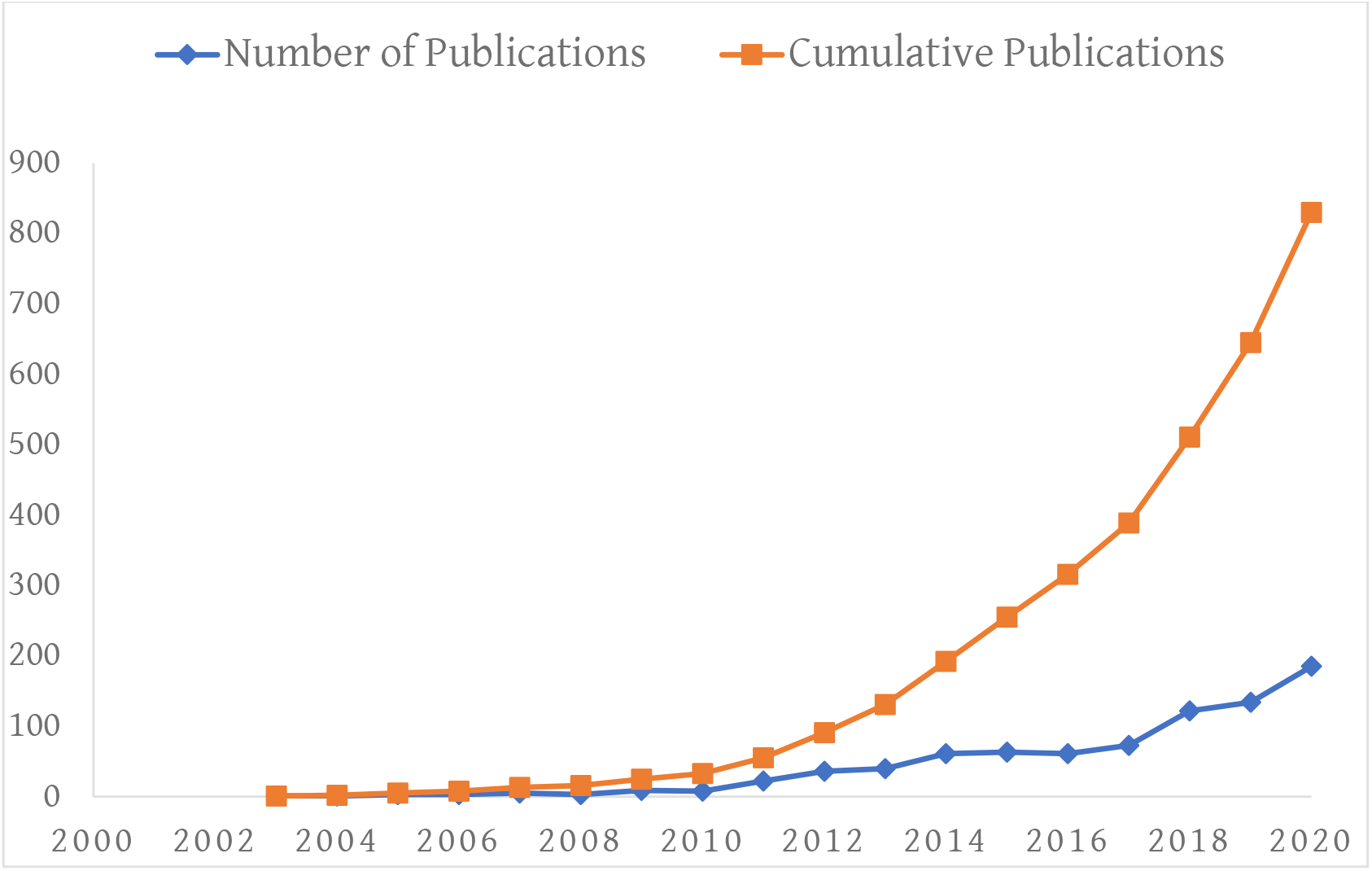
Trends of scholarly publications on syndemics.

### 3.3 Top contributors in syndemics research

Table 2 provides an overview of the top ten contributing authors who contributed to the published manuscripts on syndemics. Among the authors, Safren S.A. had a higher number of publications (n = 26), followed by Singer M (n = 24) and Halkitis P.N. (n = 17). Moreover, Singer M. had the highest number of citations (n = 949), followed by Stall R. (n = 679) and Mendenhall E. (n = 555). Singer M. also had the highest h-index (12) and g-index (24) among the top authors.

Most articles on syndemics were published in AIDS and Behavior journal (7.23%, n = 60), followed by Annals of Behavioral Medicine (3.73%, n = 31) and AIDS Care (2.89%, n = 24). Four out of the top ten journals were associated with AIDS and STI-related topics, whereas all journals were related to epidemiological, behavioral, anthropological, social, and public health sciences (Table 3).

**Table 2:** Top ten authors contributing to syndemics research.

| **Author** | **Number of Publications** | **Year of first publication** | **h-index** | **g-index** | **Total citations** |
| --- | --- | --- | --- | --- | --- |
| Safren SA | 26 | 2010 | 9 | 22 | 497 |
| Singer M | 24 | 2003 | 12 | 24 | 949 |
| Halkitis PN | 17 | 2010 | 11 | 17 | 484 |
| Mayer KH | 14 | 2015 | 6 | 14 | 229 |
| Stall R | 14 | 2005 | 11 | 14 | 679 |
| Mendenhall E | 13 | 2014 | 8 | 13 | 555 |
| O'Cleirigh C | 12 | 2015 | 6 | 12 | 200 |
| Gilbert M | 9 | 2017 | 4 | 6 | 45 |
| Pitpitan EV | 9 | 2013 | 6 | 9 | 102 |
| Reisner SL | 9 | 2009 | 8 | 9 | 349 |

**Table 3:** Top ten journals publishing syndemics research.

| Journal's name | Total publications on syndemics | Percentage | Impact factor (Journal citations reports 2019) |
| --- | --- | --- | --- |
| AIDS and Behavior | 60 | 7.22% | 3.147 |
| Annals of Behavioral Medicine | 31 | 3.73% | 4.475 |
| AIDS Care Psychological and Socio-medical Aspects of AIDS HIV | 24 | 2.89% | 1.894 |
| Archives of Sexual Behavior | 18 | 2.17% | 3.131 |
| Social Science Medicine | 17 | 2.05% | 3.616 |
| American Journal of Public Health | 16 | 1.93% | 6.464 |
| BMC Public Health | 15 | 1.81% | 2.521 |
| Global Public Health | 15 | 1.81% | 1.791 |
| Journal of Acquired Immune Deficiency Syndromes | 15 | 1.81% | 3.475 |
| Annals of Anthropological Practice | 14 | 1.69% | N/A |

### 3.4 Institutional research collaborations

Table 4 shows the top 10 institutions that were affiliated with syndemics-related research. The majority of the studies were authored by scholars from the University of California System (11.93%, n = 99), followed by Harvard University (8.19%, n = 68), and the Johns Hopkins University (6.63%, n = 55). Figure 2 shows extensive collaborations between the key affiliating institutions with a higher publication frequency (n = 5 or above), which highlights the interconnectedness of top contributing institutions in collaborative research on syndemics.

**Table 4:** Top ten institutions affiliated with syndemics-related publications.

| **Name of the institutions** | **Number of publications** | **Percentage** |
| --- | --- | --- |
| University of California System | 99 | 11.93% |
| Harvard University | 68 | 8.19% |
| Johns Hopkins University | 66 | 7.95% |
| Pennsylvania Commonwealth System of Higher Education | 55 | 6.63% |
| Yale University | 45 | 5.42% |
| University of Toronto | 43 | 5.18% |
| University of Miami | 42 | 5.06% |
| University of Pittsburgh | 38 | 4.58% |
| New York University | 37 | 4.46% |
| University of Connecticut | 35 | 4.22% |

**Figure 2:**
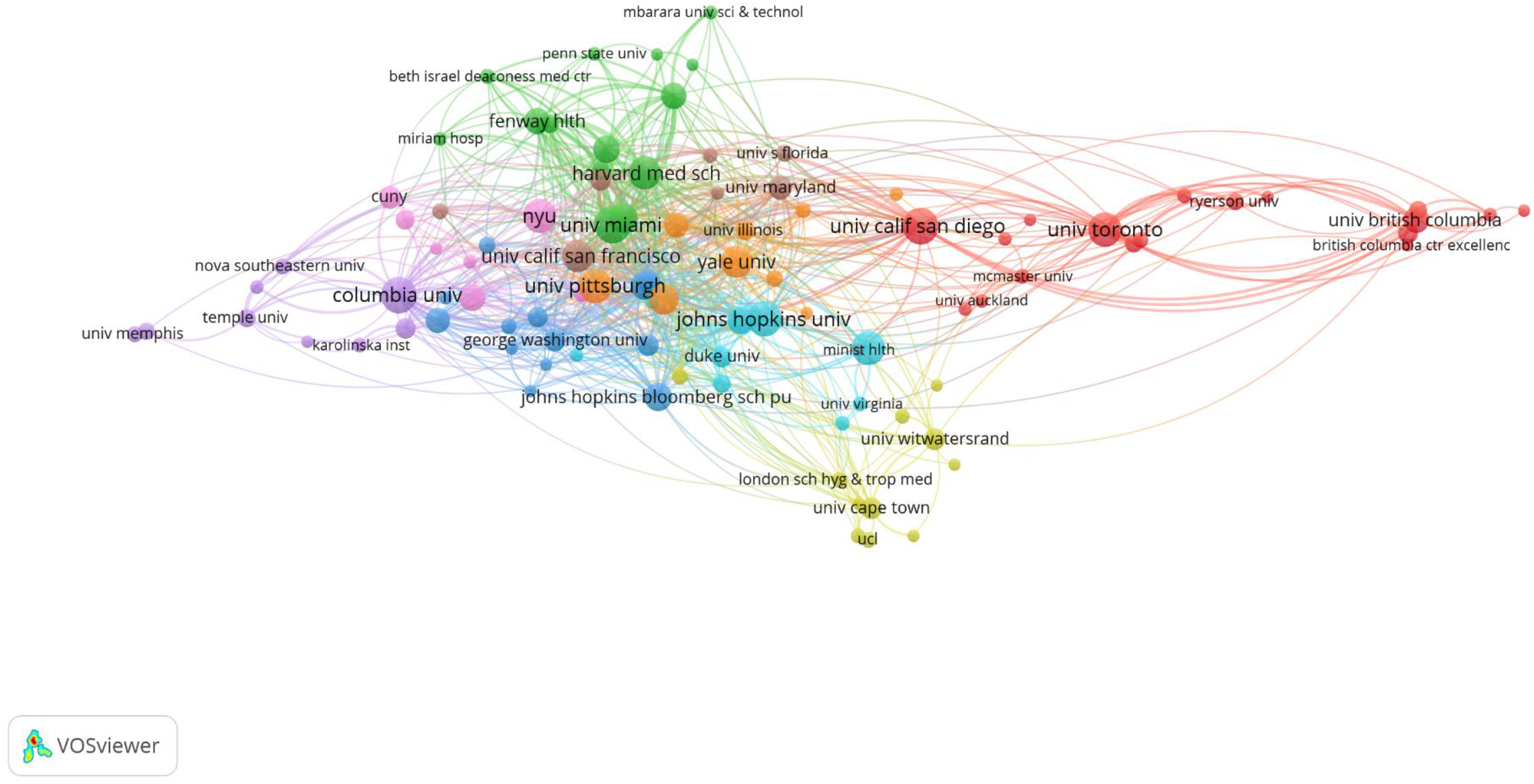
Institutional collaborations on syndemics-related research.

### 3.5 Global research collaborations

Global contributors at the country level show 76 countries that published at least one document on syndemics-related topics. Among those countries, the top 10 contributors are listed in Table 5. Most articles originated from the United States (74.46%, n = 618), followed by Canada (11.93%, n = 99) and UK (6.14%, n = 51). Figure 3 shows global collaborations among participating countries (with at least one publication), highlighting a high volume of syndemics research from high-income countries compared to low- and middle-income countries. Moreover, North American and other high-income countries had stronger collaborative ties on syndemics research that informs higher cumulative production of scientific research from these regions.

**Table 5:** Top contributing countries on syndemics research.

| Countries | Number of publications | Percentage |
| --- | --- | --- |
| USA | 618 | 74.46% |
| Canada | 99 | 11.93% |
| UK | 51 | 6.14% |
| South Africa | 44 | 5.3% |
| Australia | 28 | 3.37% |
| Peoples' Republic of China | 27 | 3.25% |
| Mexico | 20 | 2.41% |
| India | 19 | 2.29% |
| Brazil | 17 | 2.04% |
| Netherlands | 11 | 1.33% |

**Figure 3:**
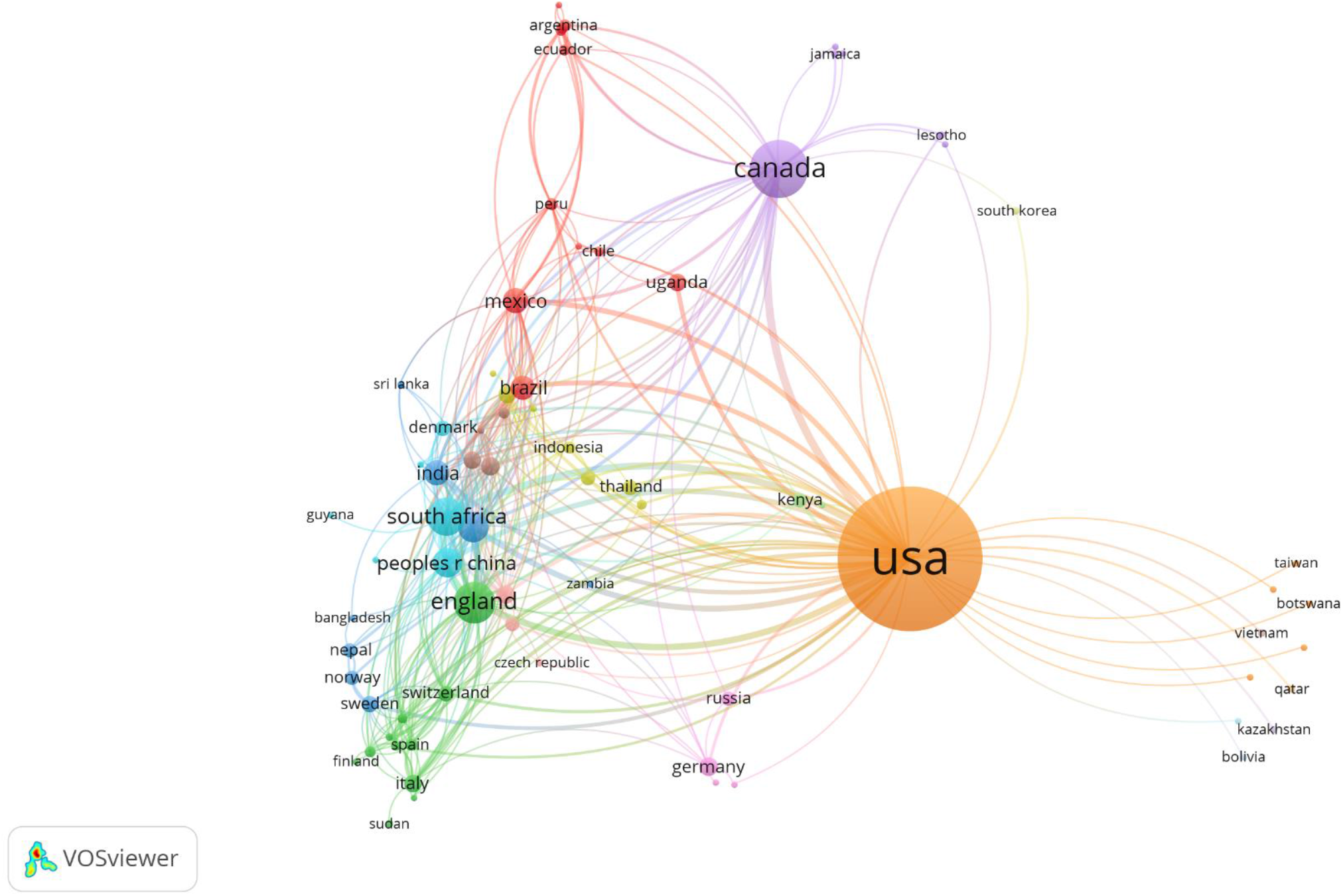
Country-level collaborations on syndemics-related research.

### 3.6 Research areas on syndemics

Several research areas were identified within the broader umbrella of syndemics using clusters of keywords that overlapped with each other. We mapped keywords that had a frequency of 5 and above across the collective bibliography. A total of 104 top keywords were identified and clustered to generate common areas of research in multiple clusters highlighted in the same color. Figure 4 shows the distribution of keyword clusters where the greater size of a circle represents a higher number of publications on that keyword and the thickness of connecting line between circles represents the intensity of coexistence of these terms in the literature.

**Figure 4:**
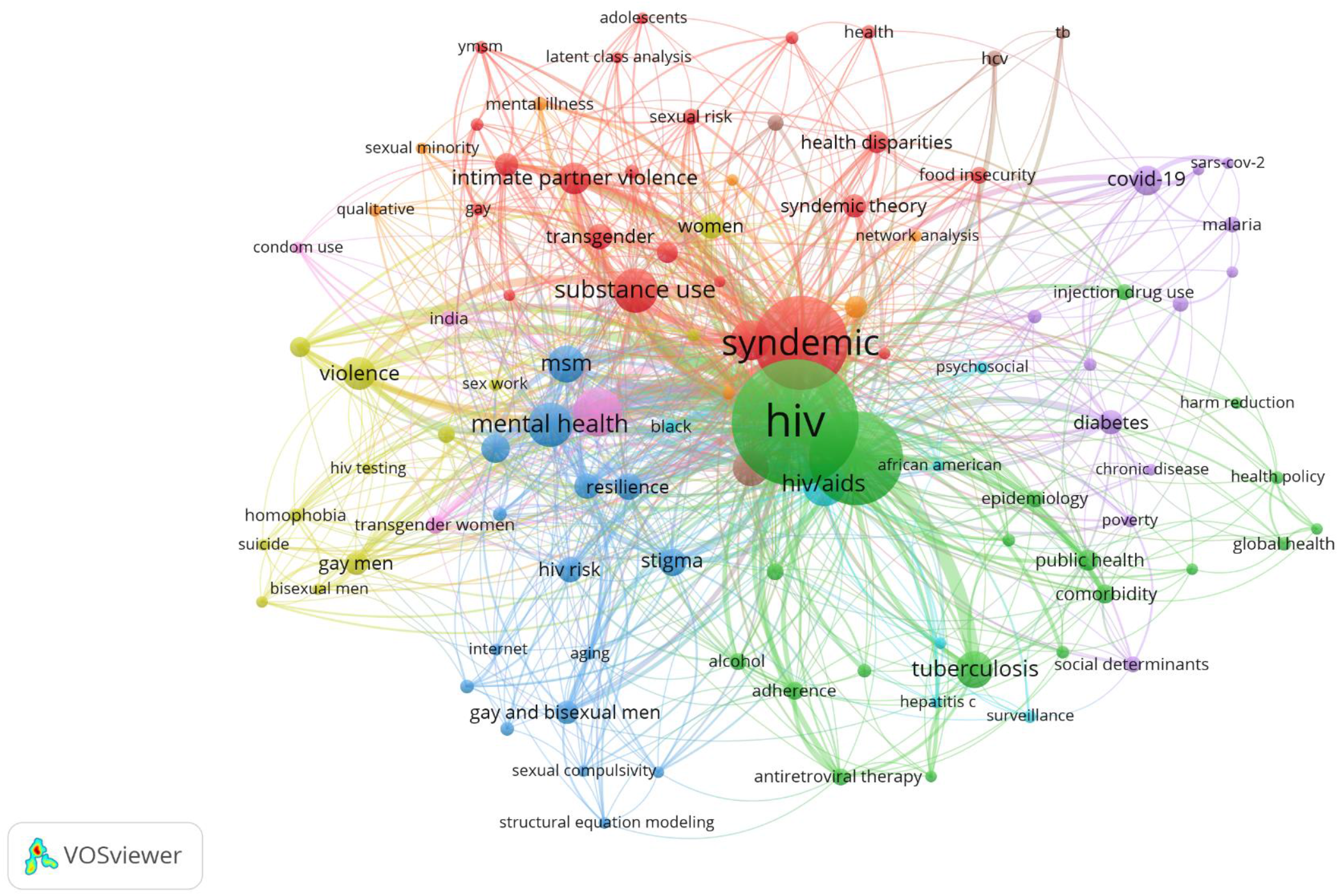
Research hotspots in syndemics research.

Amongst multiple research hotspots, the first cluster highlighted in red consisted of keywords related to syndemic theory and associated concepts such as syndemic, substance use, intimate partner violence, depression, health disparities, transgender, gay, bisexual, food insecurity, and intersectionality. Several studies were identified that used these keywords, indicating their relevance to this research cluster. For example, Couture and colleagues adopted the syndemic framework to evaluate the effects of comorbid psychosocial problems on the risk of physical and sexual violence on female entertainment and sex workers (FESW) in Cambodia [32]. They reported a high burden of client-perpetuated violence that was associated with housing insecurity, substance use, and psychological distress. FESW with two psychosocial conditions had twice the odds (adjusted odds ratio [AOR] = 2.08; 95% CI 1.00-4.31), whereas women with 5-6 psychosocial conditions had eightfold higher odds (AOR = 8.10; 95% CI 3.4-19.31) of violence, highlighting a syndemic model of cooccurring psychosocial problems. Another study examined the prevalence and correlates of trauma in South African youth living in syndemic HIV risk [33]. More than 99% of the participating youths experienced at least one potentially traumatic event (PTE), whereas a high PTE score associated with high food insecurity among adolescent men (AOR 2.63, 95% CI = 1.36-5. 09) and women (AOR = 2.57, 95% CI = 1.55-4.26, respectively). This study reported biopsychosocial pathways including depression and inconsistent condom use as pathways of syndemic of trauma and HIV in that context.

The second cluster illustrated in green includes keywords on syndemics of infectious diseases such as HIV, AIDS, tuberculosis, STIs, hepatitis c, antiretroviral therapy, adherence, comorbidity, and coinfection. For example, Dyer and colleagues conducted a cohort study among 301 men who have sex with men (MSM) to assess syndemic relationships [34]. They reported that depression symptoms were associated with sexual compulsiveness (odds ratios [OR]: 1.88, 95% CI = 1.1, 3.3) and stress (OR: 2.67, 95% CI = 1.5, 4.7); sexual compulsiveness was associated with stress (OR: 2.04, 95% CI = 1.2, 3.5); substance misuse was associated with IPV (OR: 2.57, 95% CI = 1.4, 4.8); stress was associated with depression symptoms (OR: 2.67, 95% CI = 1.5, 4.7), sexual compulsiveness (OR: 2.04, 95% CI = 1.2, 3.5) and IPV (OR: 2.84, 95% CI = 1.6, 4.9). Also, men who reported three or more syndemic constituents (three or more conditions) were engaged in high-risk sexual behavior compared to men who had two or fewer health conditions (OR: 3.46, 95% CI = 1.4-8.3). Moreover, a review by Meyer and colleagues identified 45 articles that emphasized SAVA syndemic and associated conditions such as HIV-associated risk-taking behaviors, mental health, utilization of health services and medication adherence, and a bidirectional relationship between violence and HIV/AIDS [35]. This review highlighted the complex relationships and associated outcomes of poor decision making and high-risk behavior in the context of the SAVA syndemic.

In the third cluster highlighted in blue, several keywords were identified, including mental health, MSM, sexual compulsivity, stigma, resilience, internet, and aging. Studies in this cluster focused on health behavior and syndemic relationships in gay and bisexual men. For example, a study by Parsons and colleagues examined 1,033 HIV-negative gay and bisexual men living in the U.S. and found that more than 62% of men reported having at least one syndemic condition [36]. Also, HIV-related risk behavior was associated with polydrug use, sexual compulsivity, being single, and being Latino. Moreover, the risk was highest among participants with three or more syndemic conditions. Another study from Mexico conducted by Pitpitan and colleagues found that MSMS with a high number of syndemic conditions showed an increased prevalence of sexual risk-taking [37]. Moreover, MSM who were out to more people showed a weaker association between high-risk sexual behavior and syndemic conditions suggesting outness or disclosure of same-sex preference as a resilience factor from a syndemic perspective.

A fourth cluster highlighted in yellow included keywords related to psychosocial and health behavior-related keywords such as violence, substance abuse, HIV testing, homophobia, and suicide. This research cluster emphasized the growing number of studies that examined the psychosocial health of MSM and associated syndemic conditions and relationships. For example, a study by Herrick and colleagues recruited 1551 MSM and found that different life-course predictors such as internalized homophobia and victimization were significantly associated with syndemic condition as well as psychosocial health conditions including stress, depressive symptomology, substance abuse, compulsive sexual behavior, and intimate partner violence [38]. Moreover, the authors used a nested negative binomial analysis and found that the overall life course significantly explained the variability in syndemic outcomes (chi(2) = 247.94; P<.001; df = 22). Another study by Ferlatte and colleagues examined the data from a survey of 8382 Canadian gay and bisexual men and found that suicidal ideation and attempts were associated with individual marginalization and psychosocial health problems such as mental disorders, substance use, STIs, and HIV risks [39]. In addition, individuals with three or more psychosocial problems had higher odds of experiencing suicide ideation [6.90 (5.47-8.70) times] and suicide attempts [16.29 (9.82-27.02)] compared to participants with no such problems. These relationships show the complex nature of syndemic relationships among people living under psychosocial stressors that impacts their health and wellbeing.

The fifth research cluster highlighted in purple color consisted of keywords including COVID-19, obesity, diabetes, chronic disease, poverty, and social determinants of health. This cluster emphasizes a research domain that includes noncommunicable diseases and contemporary health problems such as the COVID-19 pandemic. For example, Mendenhall and colleagues conducted a study in Kenya and found that adults with diabetes shared a complex social and medical framework associated with their health conditions. People with diabetes also had comorbid anxiety, depression, and infectious diseases such as HIV/AIDS, malaria, and tuberculosis. The authors also reported that social problems were associated with biophysical suffering, whereas women had a higher burden of psychosocial distress and somatic symptoms such as multimorbidity compared to men. People with diabetes reported not only concurrent anxiety and depression but also common infections, including malaria, tuberculosis, and HIV/AIDS. Another study from Puerto Rico found that the subaltern status negatively affected obesity rates that could be attributable to limited federal assistance for health insurance and healthier food items. Moreover, weight mismanagement and a lack of healthcare providers were amongst the psychosocial challenges that were associated with the obesity syndemic in this population. Furthermore, recent studies focused on the relevance of syndemic perspectives on coronavirus disease (COVID-19) pandemic. For example, Gutman and colleagues argued that coinfections such as malaria and other parasitic diseases with SARS-CoV-2 could result in detrimental health outcomes necessitating increased testing and disease surveillance during the COVID-19 pandemic [23]. Moreover, Perez-Escamilla and colleagues discussed the persistent effects of food and nutrition insecurity associated with poor maternal and child health outcomes that can be exacerbated during the COVID-19 pandemic [40]. Furthermore, long-standing health inequities and systemic racism are associated with multimorbidity, which may have syndemic relationships leading to adverse health outcomes in marginalized communities [41]. In this regard, Poteat and colleagues used the syndemic framework to discuss the syndemic conditions in Black Americans who experienced psychosocial stressors such as mortgage redlining, history of enslavement, political gerrymandering, lack of access to healthcare, job discrimination, and health care provider bias [42]. The authors argued that racial disparities in COVID-19 require acknowledging and addressing structural racism and determinants of these chronic disparities among the affected individuals. These articles highlight the biopsychosocial challenges and their relationships that share common determinants and affect population health, predominantly in vulnerable population groups.

Three more clusters with fewer keywords appeared in the intersections of major clusters reported above. The sixth cluster identified in pink color consisted of keywords such as men who have sex with men, condom use, transgender women, and India. Moreover, keywords such as AIDS, HCV, and TB colored in brown formed the seventh research cluster on syndemics. Lastly, scattered nodes of keywords colored in orange included HIV infection, South Africa, pregnancy, qualitative, sexual minority, network analysis, and mental illness. These clusters highlight diverse topics that overlap with other clusters and highlight the interconnectedness of the keywords as well as research topics that are common across research domains.

### 3.7 Evolution of knowledge in the field of syndemics research

Research domains within the scientific field of syndemics evolved over the years, which is highlighted in Figure 5. Since most articles were published in recent years, keywords used until 2015 highlight the scholarly themes of earlier publications on syndemics. These keywords are marked in purple, which included HIV/AIDS, substance abuse, prevention, syndemic theory, coinfection, social determinants, social inequality, internet, mortality, sexually transmitted diseases, and health policy. Moreover, keywords colored in bluish-purple show a transition of those topics being used in publications around 2016, which include syndemics, tuberculosis, obesity, harm reduction, health disparities, poverty, and aging. Furthermore, keywords used across publications during 2017-18 are presented in green color, where most keywords such as syndemic, men who have sex with men, gay and bisexual men, diabetes, domestic violence, food insecurity, global health, public health, drug use, intimate partner violence, HIV risk, testing, and prevention were identified. Lastly, keywords in yellow represent topics used in articles published around and after 2019. These recent topics included COVID-19, pandemic, social determinants of health, noncommunicable diseases, network analysis, latent class analysis, pre-exposure prophylaxis, and climate change.

**Figure 5:**
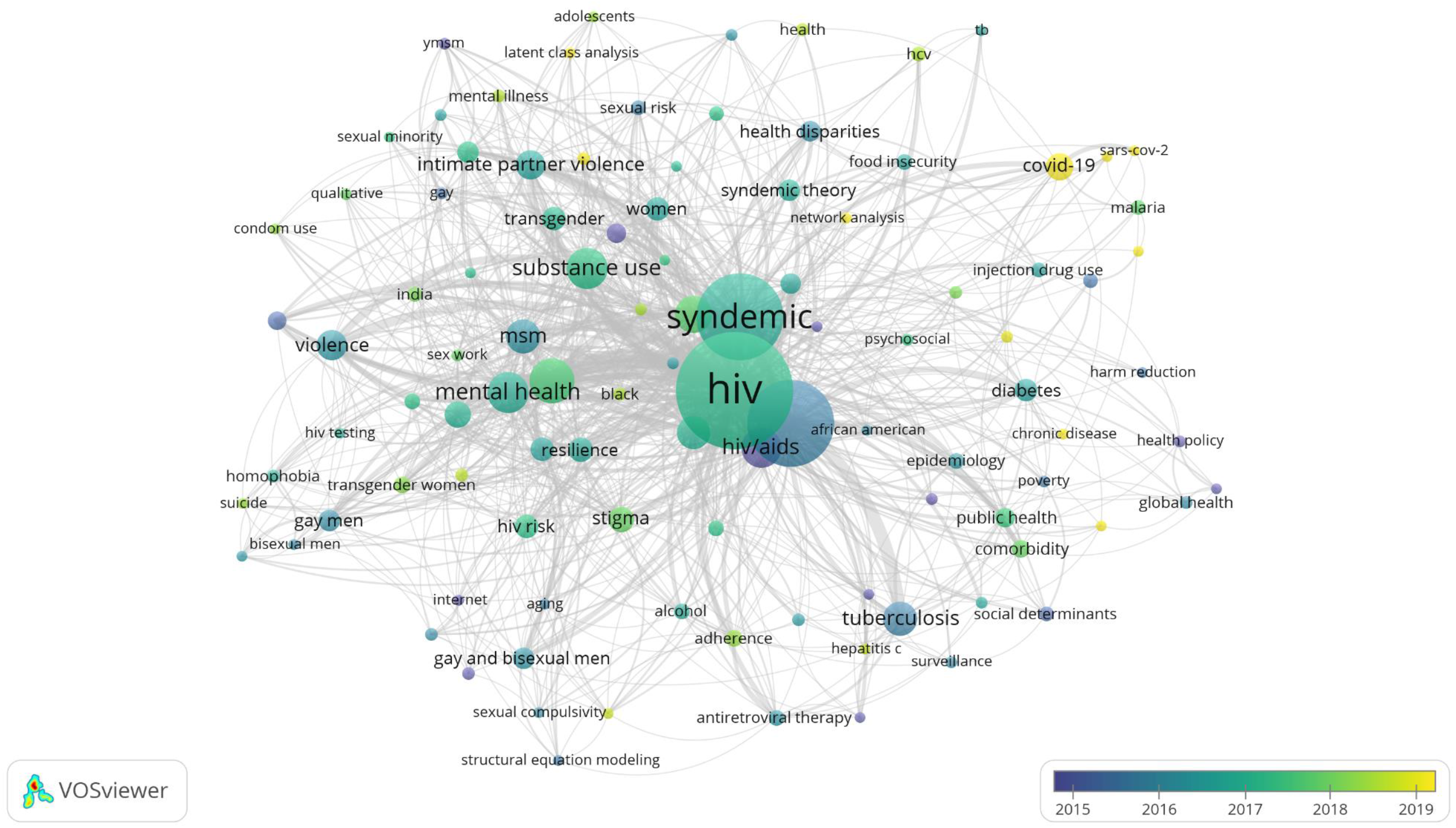
Evolution of major research topics in the field of syndemics.

An assessment of the cited references provided another perspective on how the current research publications cited and used previous scholarly items. Figure 6 represents the cited sources on the left, the most widely used keywords from the titles of cited publications in the middle, the top contributing authors on the right. Common keywords from cited sources inform the most relevant topics used from previous research, which included psychosocial health problems, gay, united states, substance use, risk, united states, prevalence, depression, health, women, and syndemics. Across these cited sources, Singer and colleagues authored five articles published from 1994 to 2017 [7, 8, 10, 12, 43]. One of earliest articles that were widely cited across syndemics literature was the article describing the development and psychometric assessment of the Center for Epidemiological Studies-Depression (CES-D) scale by Radolf, published in 1977 [44]. Furthermore, top published authors such as Singer and Stall have been contributing in syndemics research across most topics that are frequently cited across current publications.

**Figure 6:**
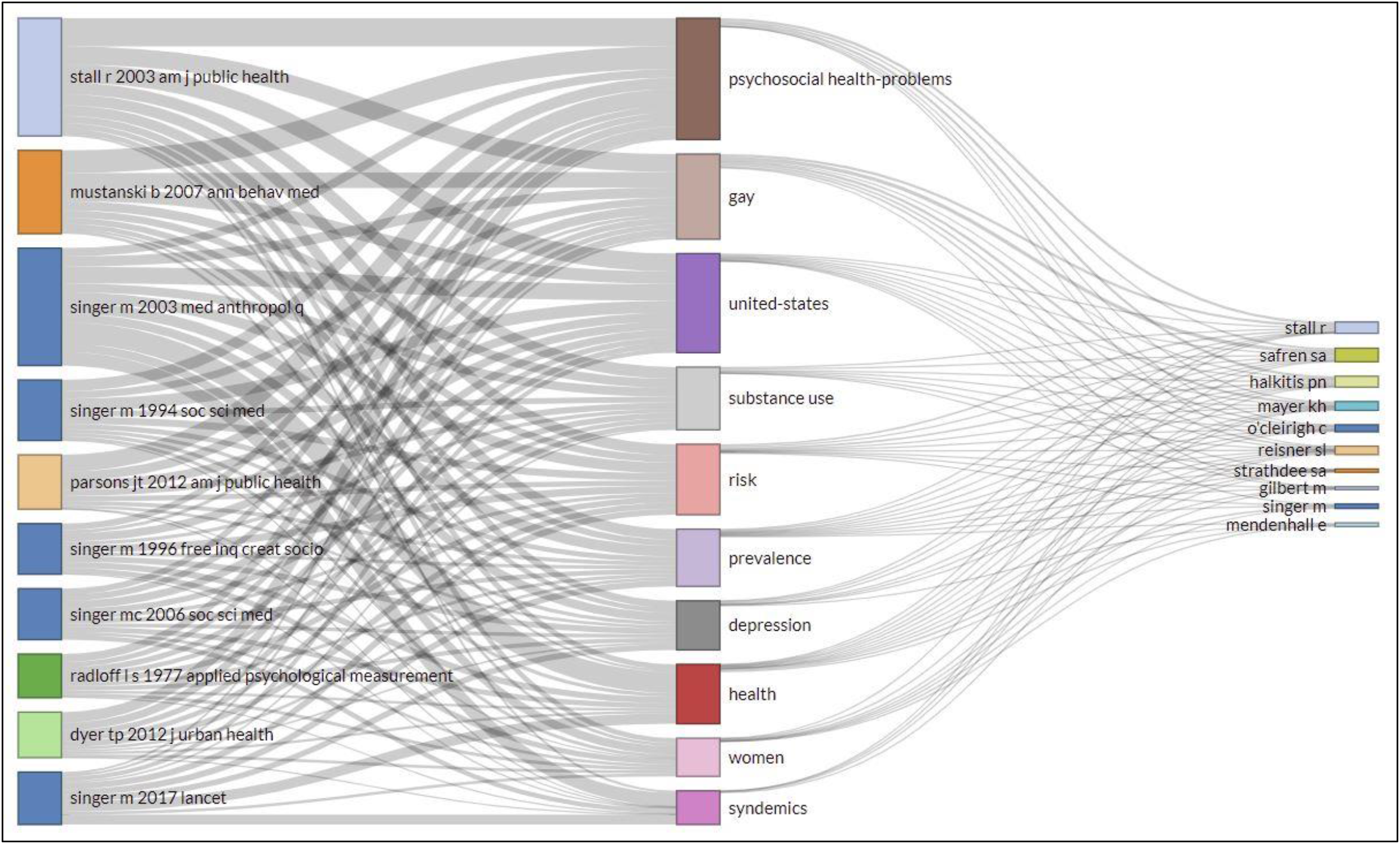
Intellectual contributions of influential articles and authors on commonly cited topics related to syndemics.

## 4. Discussion

This study evaluated the global scientific landscape of syndemics research using bibliometric measures. The findings of this study suggest a slow yet gradual increase in syndemics-related publications, which accelerated since 2013. In recent years, syndemic conditions are increasingly examined in both primary and review articles that draw intellectual inspirations from biopsychosocial literature, highlighting a transdisciplinary trend in syndemics research. Moreover, a higher proportion of original articles rather than reviews and other publication types indicates a rising body of empirical research on syndemic(s) theory and associated concepts. Furthermore, syndemics-related publications were published in general and specialty journals emphasizing the intersection of coexisting diseases, their shared determinants, and complex relationships between biosocial constructs as commonly seen in social medicine and allied fields of knowledge.

Most of the syndemics-related publications were affiliated with authors and institutions from high-income countries, whereas the U.S. had the highest contributions. In addition, heavy collaborations among authors and institutions in high-income countries indicate active research across major research entities. While some of the low- and middle-income countries (LMICs) such as Brazil, Mexico, China, South Africa, and India are engaged in syndemics research, their individual and collective contributions appeared to be limited, which informs a critical research gap in LMICs. This is consistent with previous research that reported a low scholarly output from LMICs in different areas of health sciences [45–48]. In the case of syndemics research, the historical and persistent gaps in research capacities in LMICs are likely to be compounded by the fact that syndemics have been primarily conceptualized and extensively studied by scholars and institutions in high-income countries. It is necessary to increase the research capacities in LMICs with a focus on syndemics, as those countries experience a high burden of infectious and noncommunicable diseases and poor social determinants of health. As the key concepts of syndemic(s) theory suggest [6, 7, 10], studying syndemics in such underprivileged contexts can potentially offer unique and diverse scientific perspectives on the biopsychosocial formation of health and illness in respective populations. It may enrich the current knowledge base on syndemics and inform the future transformations of medical and social care as well as disease prevention globally.

Syndemics research focused on diverse topics, among which HIV-related syndemics were extensively studied across the literature. As evident in research hotspots, HIV/AIDS and associated health behaviors and outcomes were common in more than one cluster of keywords. This frequent appearance of HIV/AIDS in the syndemics research landscape can be attributable to several factors. First, HIV/AIDS-related conditions were amongst the earliest syndemics that were conceptualized and examined in the history of the syndemic theory. For this reason, many articles focusing on non-HIV syndemics also referred to the case of previous HIV syndemics such as SAVA [9, 10]. Secondly, syndemics literature from the US and other developed countries present studies and cases of multiple health problems and their synergies within a syndemic framework [6, 9], where the study populations were urban poor from inner-city neighborhoods. As we learn from social epidemiological and anthropological studies [9, 36], biosocial challenges associated with HIV/AIDS were highly prevalent in these population groups. Therefore, the academic discourses on the social reality of health and wellbeing in these marginalized people would remain incomplete without studying HIV/AIDS and related issues. Lastly, literature from the global context, particularly from Sub-Saharan countries, highlighted the burden of HIV/AIDS in the context of the continued burden of infectious and chronic diseases in those contexts [6, 9]. For this reason, we can find studies that provide national and regional assessments of HIV/AIDS-related syndemics from those countries, which also compared their findings with what was known from similar studies conducted in developed nations. As many scholars described HIV/AIDS as a pandemic rather than just a disease outbreak [49, 50], global studies comparing and connecting the concepts and associated evidence revealed the worldwide relevance of HIV/AIDS in syndemics research.

In addition to HIV/AIDS, syndemics literature also discussed infectious diseases such as tuberculosis and COVID-19. While tuberculosis research was often studied as a comorbid condition in people living with HIV/AIDS [6, 9], recent literature examined the relevance of comorbid diseases that are relevant to susceptibility to COVID-19 and subsequent health outcomes [42, 51–53]. Given the growing burden of biopsychosocial challenges associated with COVID-19 [41, 54–59], it is necessary to investigate bio-bio and bio-social interactions between coexisting health problems in COVID-19 patients and survivors. Such research may reveal the true burden of disease clustering, biopsychosocial relationships, shared determinants, and health outcomes in the context of this pandemic. Despite a growing body of evidence that is mostly epidemiological and predominantly cross-sectional in nature, transdisciplinary and longitudinal investigations would be necessary to understand syndemic aspects of this pandemic. Furthermore, COVID-19 has demonstrated the vulnerability of people with noncommunicable diseases who are more likely to have adverse outcomes [41, 42, 53]. Limited literature exists on syndemics of non-HIV chronic diseases [9], which necessitates a more comprehensive yet inclusive study of communicable and noncommunicable diseases from a syndemic perspective.

Social determinants of health constituted a significant proportion of syndemics research. This is consistent with the fundamental idea of syndemic conditions that have common social factors and bio-social interactions [9, 10, 12]. However, there were distinct patterns in research on social factors that influenced health status, disease development, and subsequent outcomes in study populations. As evident in research hotspots, sexual and gender minorities such as men who have sex with men, bisexuals, and transgender people were frequently studied in the syndemics literature. Although many of those studies examined health problems and associated factors in the context of HIV/AIDS [6], studies have also presented their psychosocial vulnerability due to minority stressors such as social stigma and other determinants of health in the affected individuals. The added value of those studies may include, but not limit to, a broader understanding of how gender roles and norms may have biosocial interactions with pre-existing health conditions, contribute to the progression of diseases, and impact their health and wellbeing at the individual and population level. Such research findings must be translated to clinical and social decision-making addressing the stressors that affect health outcomes in gender minorities. Moreover, future syndemics research may need to extend the scope and depth of conceptual and empirical investigations on other psychosocial stressors such as racism, poverty, lack of education, unemployment, inadequate access to health and social care, xenophobia, and other means of marginalization. In recent years, public interests in those issues reflect not only their relevance to the social oppression experienced by minorities but also inform a critical need to examine the biopsychosocial dynamics of such stressors to understand how they may determine health and social outcomes in respective populations.

The current literature emphasizes diseases and their interactions among individuals in the social context. However, socioecological perspectives may inform the need for research in several under-investigated areas that may provide a more accurate understanding of how syndemics work in individuals and populations [60, 61]. For example, the role of caregivers and family members is critical for the psychosocial wellbeing of the affected individuals [48, 62, 63]. Moreover, caring for someone with one or more chronic diseases is reported to be associated with adverse health and social outcomes among family caregivers [64, 65]. Since they share the same or similar social and contextual factors relevant to the respective syndemics, future research should investigate the roles of and impacts on informal caregivers alongside primarily affected individuals in syndemic scenarios. Furthermore, syndemic conditions, as well as their determinants and consequences, may not have equal impacts on people of different age groups. Disease dynamics and associated social forces are likely to be different in children, adolescents, young adults, and older adults with varying sociodemographic characteristics [9, 66, 67]. Future research should examine syndemics in different population groups, associated biopsychosocial relationships, and multiple outcomes.

From a health services perspective, syndemics may impose unique challenges to the affected individuals requiring personalized care that may address multiple health concerns at a time. The recent technological advancements have facilitated the digitalization of health services with a focus on personalized or precision health [68–70]. Syndemics research in the digital age should explore avenues of integrating interventions that target intersections of coexisting diseases and associated factors in specific contexts. Such integrative technological measures may inform spatial and temporal challenges through real-time measurements [71, 72], and enable the practitioners to mitigate the burden of multimorbidity using evidence-based digital and traditionally delivered interventions.

Given the complex nature of syndemics, implementation research on coexisting diseases, common determinants, cumulative impacts, and shared sociocultural aspects should be conducted, and the findings should be incorporated in public health and social welfare policymaking. Despite a growing need for translating the current evidence to transform clinical and social care for syndemics, it is necessary to acknowledge that the current literature does not emphasize implementation dimensions of complex health and social problems. Researchers and decision-makers should recognize this gap as the persistence of comorbid disease would be critical for common healthcare operations such as patient-provider interactions and the delivery of health services. Institutional measures, including sensitizing the stakeholders and establishing evidence translation systems for practitioners, should be prioritized for minimizing knowledge-to-practice gaps in this regard.

Syndemics research highlights global health disparities by emphasizing the roles of context-specific forces and determinants of health [9, 13]. Therefore, local and global health systems should be examined from a syndemic perspective, which may enable health system strengthening that results in a better understanding of concurrent syndemics, future population health challenges, and how to respond to the same in a systematic way. Such insights on syndemics would also necessitate the active participation of global health organizations and member countries to use their collective resources to combat global health crises such as infectious disease outbreaks, food insecurity, climate migration, and health inequities that continue to affect populations. In these processes, the role of political commitment and collaborations will be of paramount importance.

From a health promotion perspective, it is critical to engage communities who are vulnerable to syndemics to increase their awareness on social determinants of cooccurring diseases [73, 74]. The public should be informed through targeted and mass media interventions regarding common challenges and preventive measures. However, empowering community-based organizations to assess their risks and address the same using local resources would be far more impactful. Such efforts may require technical assistance and external support, which should be organized through public health institutions. As syndemics researchers argue that context-specific factors and biopsychosocial interactions make syndemics unique to individuals and populations [9, 43, 52], participatory research and action plans may create opportunities to exchange knowledge and develop context-appropriate interventions for syndemics.

Syndemics research highlights an intersection of academic contributions from multiple scholarly disciplines, including medical anthropology, social epidemiology, health policy, health promotion, clinical sciences, and social sciences [6, 33, 61, 73]. The combined use of multiple quantitative and qualitative methods has complemented the investigations of complex syndemic problems in current literature, which couldn’t have been understood using any single-best method [6, 9, 17]. Arguably, syndemics necessitate transdisciplinary approaches to integrate multiple research methodologies to answer complex research questions. Such integrative measures may help the scholars to apprehend the ontology and phenomenology of syndemics from shared epistemological perspectives.

Global research on syndemics informs the deteriorating effects of cooccurring health problems in populations living under chronic social stressors [8, 10, 34, 60]. Understanding these complex scenarios may offer insights on humanitarian challenges that are infrequently studied and discussed in contemporary health and social policy discourses. Notably, the production of health in a population reflects its commitment to improve people’s wellbeing and maximize public welfare through regulating factors that may be harmful to individual and social health. Such initiatives may also include mandates for better access to health services irrespective of social, economic, cultural, geographic, and other differences across individuals and communities. However, many high-income countries are far behind in ensuring equitable access to health. Previous research on political determinants of health informs critical challenges such as systematic racism, implicit bias, environmental injustice, and other structural factors that share the policy responses to public health problems [75–78]. From an ethical perspective, it is a shared responsibility of healthcare providers, researchers, and organizations to advocate for using science for bringing social justice through meaningful changes in the socio-political determinants of syndemics and health disparities in marginalized populations.

## 5. Limitations

There are several limitations of this meta-knowledge study, which are necessary to understand the scope and findings of this study. Also, the limitations of this study would inform future research addressing the theoretical, methodological, and empirical shortcomings of the current findings. One such limitation is the choice of the database; although WoS is one of the most inclusive sources of bibliographic data, it may not contain all articles published on syndemics. Therefore, this study could not include potential studies that may exist elsewhere. Although this limitation is very common across knowledge mapping studies [18, 20, 24], we encourage methodologists and other scholars to continue intellectual discourses on how the global knowledge community can find opportunities to harmonize citations data across databases. This is extremely challenging as journals are not universally indexed, and databases do not contain bibliographic data on all key variables of interest. This challenge necessitates an integration of technological advancements and cooperation between database authorities to facilitate a uniform distribution of scholarly resources globally. Another challenge was the framing of syndemics in the published literature. As found in the previous synthesis of empirical research [9], many scholars do not examine all criteria of syndemics in their studies. For example, the coexistence of multiple health problems without specifying the bib-bio or bio-social relationships is often studied within the syndemic framework. Also, it is possible to describe all criteria of syndemic without using the framework, which is perhaps a major challenge leading to a limited observation of literature on complex health problems. We recommend wide scholarly communication of syndemic theory and adoption of relevant keywords and explanations whenever possible. This may eliminate the existing knowledge biases and improve future mapping of the global knowledge landscape. Lastly, despite using contemporary knowledge mapping approaches, this study may not inform population-level estimates or determinants of syndemics. As we discussed earlier, the field of syndemics research is evolving, and this study highlighted key research areas within this growing field of knowledge. We recommend further primary studies and evidence synthesis on disease clusters that have synergistic effects, interactions, and shared sociocultural factors that may inform specific insights on context-specific syndemics.

## 6. Conclusions

In this meta-knowledge study on global research on syndemics, we found a limited yet growing body of scientific literature on syndemics and associated population health problems. Most studies on syndemics were published from high-income countries, and they included diverse topics ranging from STIs to various social determinants of health. Further research is needed to understand the dynamics of multiple coexisting infectious and noncommunicable diseases across global populations with a focus on cumulative disease burden, biopsychosocial relationships, and common structural forces that are associated with health and wellbeing. The findings of this study highlight the scope of syndemics to inform advanced research, health policymaking, and practices, not only through focusing on a single disease but also addressing health inequities in the intersections of multiple health problems through inclusive, context-specific, socioculturally appropriate, and evidence-based approaches.

## Data Availability

Data on materials used in this study can be found in the following public repository (https://osf.io/b4mvr/).

## Acknowledgment

None.

